# Knowledge of and Attitudes on Artificial Intelligence in Healthcare: A Provincial Survey Study of Medical Students

**DOI:** 10.1101/2021.01.14.21249830

**Authors:** Nishila Mehta, Vinyas Harish, Krish Bilimoria, Felipe Morgado, Shiphra Ginsburg, Marcus Law, Sunit Das

## Abstract

**Background:** There has been growing acknowledgement that undergraduate medical education (UME) must play a formal role in instructing future physicians on the promises and limitations of artificial intelligence (AI), as these tools are integrated into medical practice.

**Methods:** We conducted an exploratory survey of medical students’ knowledge of AI, perceptions on the role of AI in medicine, and preferences surrounding the integration of AI competencies into medical education. The survey was completed by 321 medical students (13.4% response rate) at four medical schools in Ontario.

**Results:** Medical students are generally optimistic regarding AI’s capabilities to carry out a variety of healthcare functions, from clinical to administrative, with reservations about specific task types such as personal counselling and empathetic care. They believe AI will raise novel ethical and social challenges. Students are concerned about how AI will affect the medical job market, with 25% responding that it was actively impacting their choice of specialty. Students agree that medical education must do more to prepare them for the impact of AI in medicine (79%), and the majority (68%) believe that this training should begin at the UME level.

**Conclusions:** Medical students expect AI will be widely integrated into healthcare and are enthusiastic to obtain AI competencies in undergraduate medical education.

## INTRODUCTION

Many experts have discussed the anticipated role of artificial intelligence (AI) in medicine as tools to improve medicial decision-making and efficiency (Rajkomar, Dean and Kohane, 2019; Topol, 2019). Much of this discussion has been grounded in recent improvements in the performance of AI algorithms and the growing availability of large medical datasets to test these algorithms (Oakden-Rayner, 2020). As AI is introduced and eventually integrated into healthcare, physicians will be expected to use AI tools to monitor, diagnose, and treat their patients.

There is also a growing acknowledgement that undergraduate medical education (UME) must play a formal role in instructing future physicians on the promises and limitations of AI tools in medical practice (Masters, 2019; Davenport and Kalakota, 2019; Paranjape *et al*., 2019; Kolachalama and Garg, 2018; Wartman and Combs, 2019; McCoy *et al*., 2020). By the time today’s medical students complete their postgraduate training—in many cases almost a decade later—it is expected that AI tools in medicine will be more commonly integrated into clinical decision-making workflows (National Health Service, 2019). UME provides a key opportunity to give future physicians a broad understanding of the capabilities, limitations, and consequences of the tools they will use in their training and beyond.

Looking abroad, the United Kingdom’s National Health Service recommended the integration of AI competencies in every level of healthcare professional training (National Health Service, 2019). In the United States, the American Medical Association recently adopted a policy promoting enhanced training in AI across the continuum of medical education (American Medical Association Board Report, 2018). Similarly, there has been significant Canadian student interest in AI competencies in undergraduate medical education (Bilimoria *et al*., 2019). The desire to introduce AI into medical education across students, health services, and professional organization highlights the need for further exploration on how such a curriculum should be developed.

The Royal College of Physicians and Surgeons of Canada Task Force on AI and Emerging Digital Technologies recently released a report on the expected impacts of AI on specialty fellowship training, which recommended the development of guidelines for integrating instruction on AI across residency training programs (Royal College of Physicians and Surgeons of Canada, 2020). No such efforts currently exist for UME programs. Further, while several commentaries have published calls to action to integrate AI into medical education and have described broadly what key competencies could be, there remains a paucity of competency frameworks or learning objectives to guide the creation of AI in medicine curricula at the UME level (Masters, 2019; Davenport and Kalakota, 2019; Paranjape *et al*., 2019; Kolachalama and Garg, 2018; Wartman and Combs, 2019; McCoy *et al*., 2020).

The development of novel medical school curricula involves a collaborative effort among subject matter experts (e.g. computer scientists, data scientists) and specialists in medical pedagogy. Best practices in curriculum development encourage targeted needs assessments among medical learners (Thomas *et al*., 2016). As the momentum to integrate AI competencies into medical education builds, understanding medical students’ baseline knowledge, perceptions, and concerns about clinical AI may provide further insight into priority areas for curriculum development. Thus far, studies surveying medical students on this topic have focused principally on the perceived impact of AI on radiology (Gong *et al*., 2019; Sit *et al*., 2020; Pinto Dos Santos *et al*., 2019). To develop educational content relevant for all medical students, however, a broader assessment of student perceptions should be undertaken. To this end, we administered a survey to students across four medical schools in Ontario. This survey sought to serve as an exploratory analysis of medical students’ knowledge of AI, perceptions on the role of AI in medicine, and preferences surrounding the integration of AI competencies into medical education.

## METHODS

### Survey Design

We designed an online survey using SurveyMonkey. A survey was conducted rather than interviews or focus groups as our goal was increased representation (number of students), and to capture standardized data on individual student knowledge and perceptions on a wide variety of aspects on AI in medicine, and as opposed to an in-depth exploration of student perspectives with a limited number of questions (Kelley *et al*., 2003). Survey questions were developed following a review of articles proposing to integrate AI competencies into UME (Masters, 2019; Davenport and Kalakota, 2019; Paranjape *et al*., 2019; Kolachalama and Garg, 2018; Wartman and Combs, 2019; McCoy *et al*., 2020), previous surveys of physician and medical student attitudes on AI in medicine (Laï, Brian and Mamzer, 2020; Blease *et al*., 2019; Oh *et al*., 2019; Blease *et al*., 2018), and consultation with educators involved in developing AI in medicine curricula at the University of Toronto Medical School. The complete survey questionnaire is included in Supplementary Figure 1.

The survey consisted of three sections. The first survey section captured demographic data, students’ education before medical school, and past exposure to AI-related content (e.g., curricular, self-directed learning etc.). The next section focused on medical students’ knowledge of AI by asking for their level of confidence in their understanding of terminologies such as AI, machine learning, neural networks, and deep learning.

The final section gathered student perceptions of AI in medicine. At the start of this section, we presented the following definition of AI to allow students unfamiliar with the term to answer questions on their perception of this topic: *AI is the theory and development of computer systems able to perform tasks that normally require human intelligence, such as visual perception, speech recognition, decision-making, and translation between languages*. The first set of questions assessed students’ beliefs about the capability of AI to eventually perform a specific task (e.g., updating medical records) at a level equivalent to a human physician. The questions in this section were based on the USAID framework for AI use cases in healthcare sections on individual health, population health, and health systems (United States Agency for International Development, 2019). This section asked students for their estimate on the likelihood of AI ever being able to replace a physician in the performance of a specific task. For those students who selected likely or extremely likely, adaptive questioning was used to gauge the timeframe in which students believed these capabilities would be achieved (Blease *et al*., 2018). The remainder of this section assessed students’ perceived impact of AI on physician job prospects, novel ethical and societal challenges, and finally, perceptions around how AI should be incorporated into medical education. At the end of this section, a final question invited students to leave any other free text comments they had on AI and medicine.

Before survey dissemination, usability and technical functionality were tested by the research team and five medical students who were not involved in the study design. No changes were made to the survey following pilot testing.

### Recruitment Process

The study was approved by the University of Toronto Health Sciences Research Ethics Board. We sent the survey to MD program staff at all six Ontario medical schools in Ontario and received responses from four schools. Medical students of all years were eligible to participate.

At three of the four medical schools that agreed to participate, the survey was disseminated via email by the administrative office of the MD programs to the MD student body. At one medical school, the survey was sent out in a newsletter to the MD student body. At all schools, the survey was open for four weeks, with a reminder email sent two weeks after the first email. Participation was voluntary and was not related to the students’ ongoing curricular activities. Students were offered entry into a gift card raffle for completing the survey. Consent for study participation was obtained through the first page of the survey, and respondent anonymity was guaranteed by design. All data were collected between February and June 2020. Multiple entries by the same respondent were mitigated through SurveyMonkey settings, which prevented the survey from being completed multiple times on the same IP address.

### Data Analysis

All completed and partially completed questionnaires were included in the final analysis. Results were exported from the SurveyMonkey website to local spreadsheets and analyzed using Microsoft Excel. Descriptive statistics on survey questions were completed, as survey results were quantified as a percentage of total respondents. Free text responses were analyzed using an inductive approach to thematic analysis (Kiger and Varpio, 2020). Two authors (NM and VH) independently identified initial codes, and then collaboratively constructed themes from the codes. We resolved any disagreements through discussion.

## RESULTS

321 medical students initiated the survey (overall response rate, 13.4%), and the completion rate of those who started the survey was 89.7% (n = 288).

### Demographics & Participant Background

Respondents were most commonly pre-clerkship (year 1 or 2) students (55.5%, n=177/319), and female (63.0%, n=201/319). The most common educational background was in biomedical and health science (88.7%, n=278/315), with only 5.4% (n=17/315) students reporting background in engineering, math, or computer science. Very few (9.4%, n=30/318) students reported formal training in computer science at the undergraduate level or higher; however, a much higher proportion of students reported the completion of self-directed learning in the form of attending a talk/lecture on AI (39.8%, n=126/317) or other training in programming/coding (26.8%, n=85/317). Complete demographic and background information can be found in Table 1.

**Table 1:** Respondent demographics and background.

| Descriptor | N (%) |
| --- | --- |
| ***School***<br>University of Toronto<br>Northern Ontario School of Medicine<br>Western University<br>Queen's University | ***Response N: 317/321***<br>165 (52.1)<br>20 (6.3)<br>107 (33.8)<br>25 (7.9) |
| ***Year of Study***<br>Year 1 (pre-clerkship)<br>Year 2 (pre-clerkship)<br>Year 3 (clerkship)<br>Year 4 (clerkship)<br>Other | ***Response N: 319/321***<br>78 (24.5)<br>99 (31.0)<br>63 (19.8)<br>74 (23.2)<br>5 (1.57) |
| ***Gender***<br>Female<br>Male<br>Non-binary / Other | ***Response N: 319/321***<br>201 (63.0)<br>117 (36.7)<br>0 (0) |
| Prefer not to say | 1 (0.3) |
| ***Highest Level of Education Before MD***<br>Bachelor's<br>Master's<br>Doctorate | ***Response N: 321/321***<br>137 (42.7)<br>118 (36.7)<br>66 (20.6) |
| ***Educational Background (Undergraduate Studies)***<br>Biomedical Science and Health Science<br>Arts and Humanities<br>Business<br>Engineering, Math, and Computer Science<br>Other | ***Response N: 315/321***<br>287 (91.0)<br>9 (2.8)<br>2 (0.6)<br>17 (5.4)<br>1 (0.3) |
| ***Undergraduate or higher training in computer science***<br>Yes<br>No | ***Response N: 318/321***<br>30 (9.4)<br>288 (90.6) |
| ***Completed any course where AI/ML/DL were taught or discussed\****<br>Yes<br>No | ***Response N: 318/321***<br>31 (9.8)<br>287 (90.2) |
| ***Attended or viewed any talks or lectures on AI***<br>Yes<br>No | ***Response N: 317/321***<br>126 (39.8)<br>191 (60.2) |
| ***Any other training in computing programming/coding***<br>Yes<br>No | ***Response N: 317/321***<br>85 (26.8)<br>232 (73.2) |

\*Includes degree courses, certificate programs, and online courses.

### Existing understanding of Artificial Intelligence

Students’ understanding of general terms was stronger than more specific terms - 83.3% (260/312) of students ‘agreed’ or ‘strongly agreed’ with the phrase “I understand what the term artificial intelligence means”, 65.9% (205/311) when asked the same about ‘machine learning’, 42.3% (133/311) for ‘neural network’, and just 18.7% (58/311) for ‘deep learning’.

### Perceptions around AI Capabilities

Students’ perceptions of AI’s potential capability in the domains of individual health, health systems, and population health are described in Supplementary Table 1. Perceptions regarding the timeline in which these capabilities will be achieved are described in Supplementary Table 2. In the narrative summary below, responses of both ‘extremely likely’ and ‘likely’ have been combined and represented as ‘likely’, and ‘extremely unlikely’ and ‘unlikely’ have been combined as ‘unlikely’.

**Table 2:**
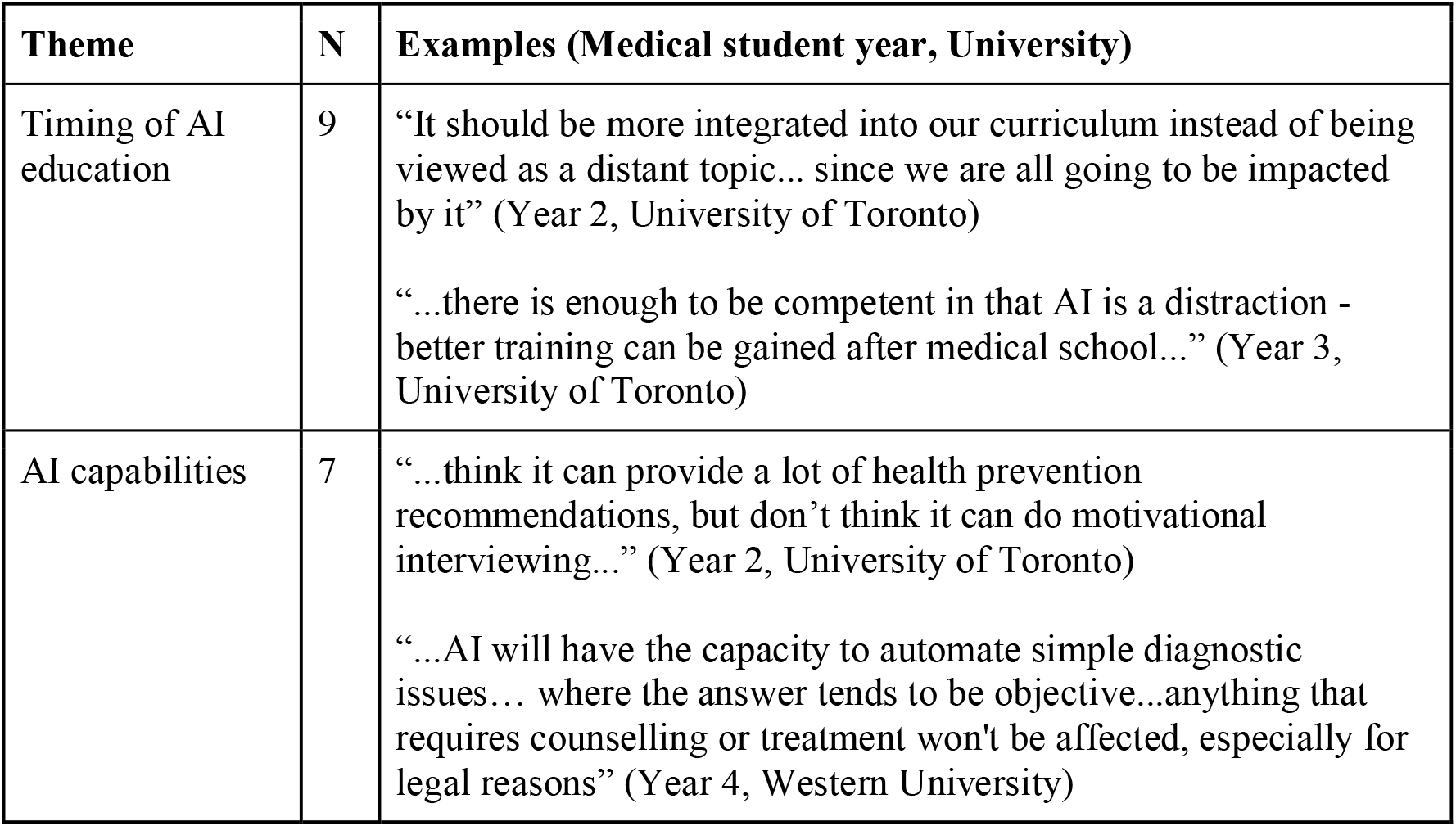
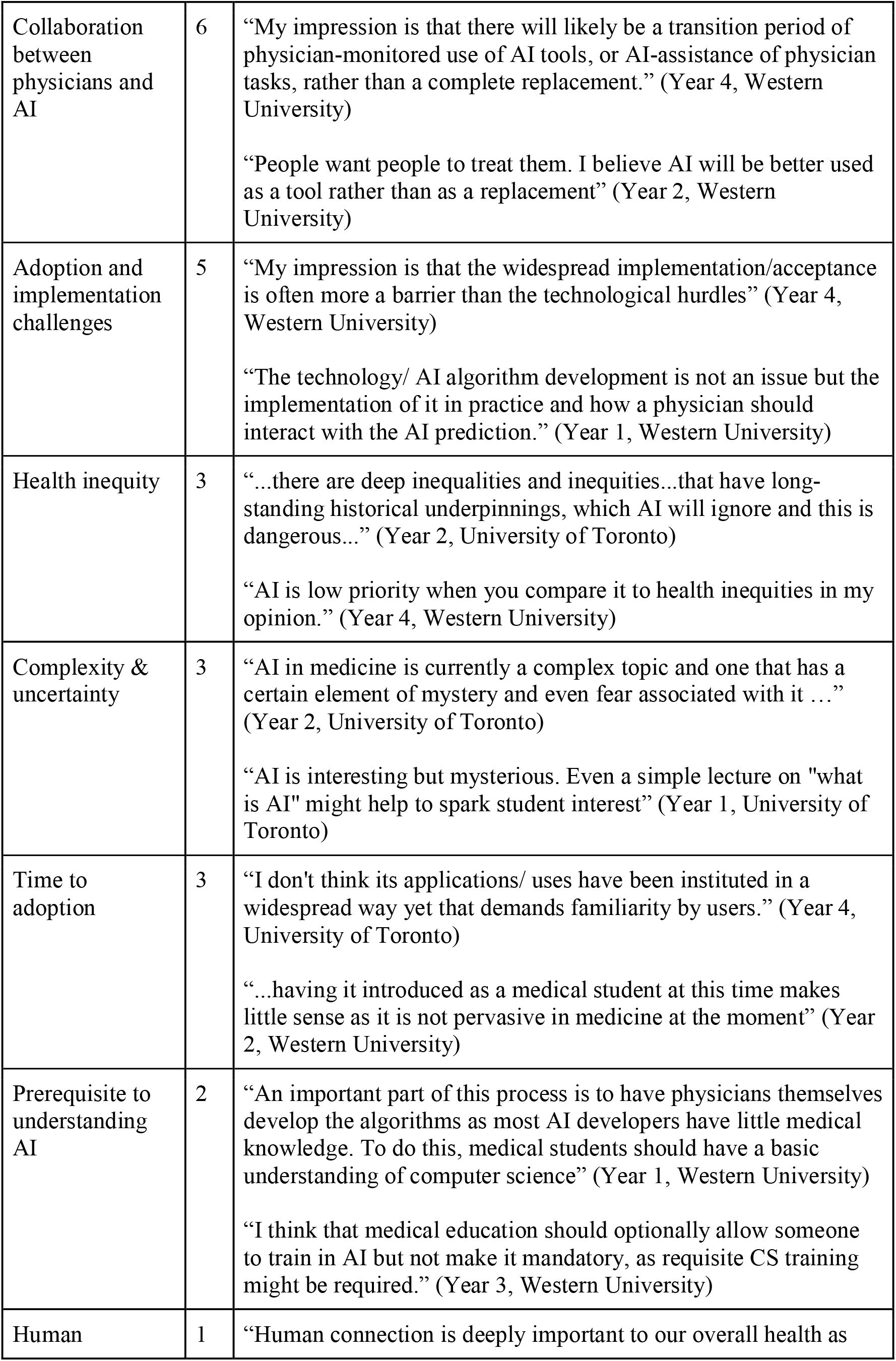
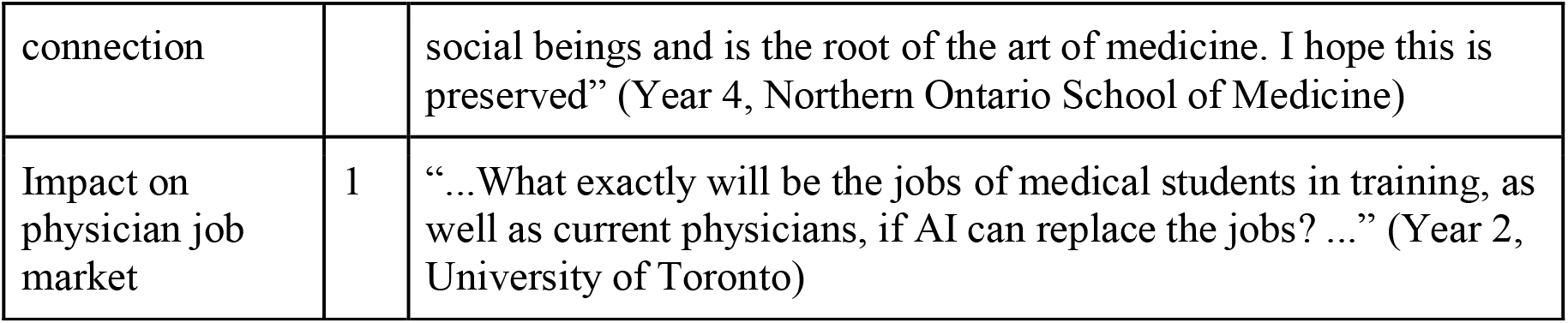
Themes constructed from free-text responses and corresponding examples.

#### Individual health

Students were generally optimistic regarding AI’s capabilities in clinical decision-making, with 71% (213/297) responding that it was likely that AI would be able to use patient information to reach diagnoses, 30% of whom believed this capacity would be attained in 5-10 years and 44% within 11-25 years. Similarly, 76% (226/296) of students responded that it was likely AI would be able to establish prognosis, with 30% feeling this would be a reality within 5-10 years, and 44% within 11-25 years. When asked about AI’s capacity to read and interpret diagnostic imaging, the vast majority of students (83%, 243/292) responded that it was likely, with 92% of these students believing this would be attained within 25 years. Many students responded that AI would be able to formulate personalized medication prescriptions—53% (156/292) felt it was likely, and the majority of those students (84%) felt this was possible in 25 years. Students were somewhat confident in AI’s role in performing robotic surgery (54% likely, 156/289), and among these students, 67% responded that it was possible within 25 years.

There were other aspects of patient care that students treated with more skepticism. Students had mixed opinions on whether AI would be able to formulate personalized treatment plans for patients, with only 29% (85/292) responding that it was likely. Students overwhelmingly responded that it is unlikely that AI will be able to provide empathetic care to patients (78%, 229/292), and even among the minority who responded that this was likely (7%, 19/292), about half felt it would take over 25 years for this to be achieved. Students responded similarly about AI’s ability to provide psychiatric/personal counselling, with 77% (223/290) reporting that this was unlikely ever to be achieved.

#### Health systems

Students expect that AI will play an integral role in health systems in the near future. The vast majority of students believed that it is likely that AI will be capable of providing documentation such as updated medical records about patients (88%, 254/289), and among those 70% responded that it would occur in the next ten years. Again, the vast majority of students (73%, 209/288) responded that it is likely that AI will assist hospitals in capacity planning and human resource management, with close to half of these students (54%) believing that this will happen in the next 10 years. Finally, similar sentiments were expressed when posed with the statement that AI will “provide recommendations for quality improvement in practices/hospitals”, with 65% (188/289) of students responding that this is likely. Again, the majority (61%) believed that this would occur in 5 to 10 years.

#### Population health

There was optimism that AI will make strides in the field of population health in the near future. The vast majority (80%, 230/289) felt it was likely that AI will be able to conduct population health surveillance and outbreak prevention. Of those students, 38.5% believed that this would happen in the next 5 to 10 years. Conversely, less than half (40%, 116/289) of students believed that AI is likely to be capable of selecting the best population health interventions; however, among these students, they mostly believe that this will occur in the next ten years (59%).

In the below three sections, unless otherwise specified, responses of ‘strongly agree’ and ‘agree’ have been collapsed into ‘agree’, and ‘strongly disagree’ and ‘disagree’ collapsed into ‘disagree’, with detailed data available in Supplementary Table 3.

### Impact of Artificial Intelligence on Profession

While students believe that AI will notably alter the job market in medicine, they are split as to whether or how this will personally affect them. When presented with the prompt, “Artificial intelligence will reduce the number of jobs available to physicians”, few students expressed strong agreement (5%, 15/288) or strong disagreement (2%, 6/288). Many students agreed (34%, 98/288), while 34% neither agreed nor disagreed. When asked if “Artificial Intelligence will reduce the number of jobs in certain medical specialties more than others”, the vast majority (87%, 250/288) of students agreed. Finally, opinions were split on whether “Artificial Intelligence will/already did impact my choice of specialty selection”: 58% of students (168/288) disagreed and 25% of students agreed (71/288).

### New Challenges Raised by Artificial Intelligence

Students are conscious of the ethical and social implications of the application of AI to healthcare but do not believe that the Canadian healthcare system is currently equipped to deal with these challenges. The vast majority of students agreed (98%, 281/288) that “AI in medicine will raise new ethical challenges”. Likewise, 95% (275/288) agreed that “AI in medicine will raise new social challenges”. This trend continued when students were posed with the statement “AI in medicine will raise new challenges around health equity” (78%, 225/288 agreed). Yet when posed with the statement, “The Canadian healthcare system is currently well prepared to deal with challenges having to do with AI”, 63% (181/288) of students disagreed, and 33% neither agreed or disagreed.

### Artificial Intelligence and Medical Education

The vast majority of students (79%, 228/288) disagreed that their medical education is adequately preparing them to work alongside AI tools. When asked whether medical training should include AI competencies, 72% (207/287) of students agreed, yet only about 52% of students agreed that this training should be mandatory. When asked when training in AI competencies should begin, the majority (68%, 195/286) selected medical school, 20% (57/286) selected residency, and 1% (3/286) selected as practicing physicians. Eleven percent (31/286) felt no training in AI competencies was necessary.

### Analysis of Free Text Responses

There were 26 free-text responses. The major themes identified were the following: when to introduce AI competencies into medical education (n= 9), the capabilities of AI in healthcare (n= 7), collaboration between physicians and AI (n= 6), and AI adoption and implementation challenges (n= 5). All themes with examples are detailed in Table 2.

## DISCUSSION

In this survey of Canadian medical students, we found that students were generally optimistic regarding AI’s capabilities to carry out a variety of healthcare functions, from clinical to administrative, but had reservations about specific task types. Students were also concerned about how AI will affect the medical job market. They believe AI will raise ethical and social implications yet are unconvinced that our health system is equipped to deal with these novel challenges. Overall, students agree that medical education must do more to prepare students for the impact of AI in medicine. To our knowledge, this is the first study assessing medical students’ knowledge and perceptions of AI in medicine broadly, providing novel insights for medical educators to draw upon in designing curriculum.

### Students’ Perceptions of the Future of Medicine and the Medical Profession

One of the main findings was that student perceptions of the future capabilities of AI showed greater confidence in AI’s ability to perform ‘objective’ tasks such as diagnosis, prognosis, interpreting imaging and formulating prescriptions, compared to tasks requiring more person-centered skills, such as psychiatric and personal counselling, or providing empathetic care. These perceptions broadly align with where AI tools have demonstrated success in research settings, such as image interpretation, diagnostics, clinical outcome prediction, and automating administrative tasks (Topol, 2019). An example of discordance between student perceptions and demonstrated AI capabilities was the use of AI-enabled tools in mental health care (i.e. conversational agents, which some patients have been shown to prefer when discussing intimate or sensitive topics) (Luxton, 2016). Students’ qualitative responses further supported the notion that AI would not be able to replace physicians in the ‘art of caring’ (Johnston, 2018). As such, these students felt that AI would be a tool used by physicians to improve their productivity, instead of entirely replacing them. This is consistent with perceptions held by key thought leaders and experts in the field of medical AI (Israni and Verghese, 2019; Verghese, Shah and Harrington, 2018).

Students’ perceptions of AI’s capabilities resonate with their opinions on how AI will impact the medical professional landscape, as they broadly expressed the belief that some specialties will be affected more than others. Interestingly, we found these perceptions were actively impacting career planning for some students: a quarter of students agreed that AI had either already or would impact their choice of medical specialty. While our survey did not probe for which specialties the threat of AI made less desirable, a 2019 study assessing Canadian medical students’ preferences for radiology found that 68% agreed that AI would reduce demand for radiologists and one-sixth of students interested in radiology would not consider it because of anxiety related to AI (Gong *et al*., 2019). A survey completed among students in the United Kingdom also found that approximately half of the surveyed students were less likely to consider radiology as a specialty due to their concerns of the impact of AI on the practice of radiology (Sit *et al*., 2020). While similar studies have not been done for other specialty areas where AI has made substantial advances (e.g. dermatology, pathology), it may be that student perceptions of AI’s emerging capabilities in these areas correlate with their concerns about decreased job demand.

### Implications for Medical Education

While there was agreement on the need to integrate AI competencies into medical training, there were conflicting opinions on when and how this should be done. Most students (68%) supported calls to integrate AI education into UME. However, a fifth felt that residency or independent practice was a more appropriate time for this education, suggesting that AI training was perceived as best delivered in a specialty-specific manner. Experts in the field of medical AI support the notion that almost every type of clinician will be impacted by AI in the future, given the breadth of existing and emerging AI applications (Rajkomar, Dean and Kohane, 2019). However, students’ perceptions highlight a desire to distinguish between underlying ‘foundations’ of medical AI to be integrated into UME, and more specialized skills to be acquired in residency or practice.

Academic discussion on the curricular content for AI education in medical schools has been limited to expert commentary (Masters, 2019; Davenport and Kalakota, 2019; Paranjape *et al*., 2019; Kolachalama and Garg, 2018; Wartman and Combs, 2019; McCoy *et al*., 2020). These commentaries agree that the goal of AI education in undergraduate medical training should be to establish a foundation in medical AI that equips graduates with a basic understanding of the technologies behind AI, their limitations, and relevant ethical and legal implications. They also share the recognition that majority of medical students may not need as much content knowledge as those who wish to be actively involved in developing and spearheading AI related innovations, and that the latter should have opportunities to further their knowledge via extracurricular, dual-degree, or certificate programs which enable acquisition of greater expertise (Paranjape *et al*., 2019; Kolachalama and Garg, 2018; Wartman and Combs, 2019; McCoy *et al*., 2020). Our survey findings reinforce these proposals. There was a keen interest among students in acquiring foundational AI competencies starting in UME. Approximately half of the surveyed students felt that AI training should not be mandatory, demonstrating an appetite for choice in how much AI-related training they engage with.

Our survey revealed areas that are currently not well addressed in the proposed AI curricula. First is the need to address AI’s impact on medical professional roles and career prospects. Our survey found that uncertainties about AI’s asymmetric impact on different specialties were already actively impacting student career planning. As medical students balance several competing factors, such as a more competitive job market, socioeconomic factors, and lifestyle when picking their specialty, AI now presents another set of considerations. Moreover, thought leaders hypothesize the evolution of certain specialties over time, such as radiology and pathology, fusing into a single ‘information specialist’ (Jha and Topol, 2016). While educators may not have anticipated that AI would be a career consideration at this time, our results indicate these discussions represent a necessary and urgent addition to UME curricula.

Furthermore, AI’s impact on health equity is a big concern amongst scholars studying clinical AI’s implications, and there may be a need to emphasize this in newly developed medical curricula. Notable examples include systemic bias identified in algorithms trained on predominately light skin-coloured patient in the assessment of dermatological findings, and challenges around applying AI to diseases and conditions relevant to resource-poor settings due to inadequate data (Gomolin *et al*., 2020; Wahl *et al*., 2018). Our results revealed near-unanimous agreement that AI would raise new ethical and social challenges. Still, fewer students were convinced that it would pose challenges around health equity (78% agreed, 16% neither agreed nor disagreed). These results suggest that students could benefit from a more nuanced discussion of what, precisely, these ethical and societal challenges are, including how they can manifest as health equity issues.

## LIMITATIONS AND FUTURE DIRECTIONS

One limitation of this study is that survey participants were from medical schools in the province of Ontario and may not be generalizable to a larger population. These four schools however, are diverse in terms of their geography (urban and rural), proximity to major academic centers, and class size, and have different curricular offerings, mitigating significant biases in the survey sample. Furthermore, generazability is limited by our response rate of 13.4%. However, the respodant pool was representative in terms of gender-63% of participants self-identified as female, consistent with estimates of the demographic makeup of Canadian medical students today (63% female), and close to that of of Canadian medical residents (53% female) (Khan *et al*., 2019; Canadian Post-M.D. Education Registry, 2018). Participation was also consistent across all four years of medical school (Year 1: 24.5%, Year 2: 31.0%, Year 3: 19.8%, Year 4: 23.2%).

Overall, the goal of this work was not to draw representative conclusions regarding all Canadian medical students, but rather to understand how medical student knowledge and perceptions could inform curriculum development, which we feel was achieved with our sample. As a next step, experts in medical AI and medical educators should synthesize proposed AI-competencies in UME curricula with our and other data of student perceptions to construct curricular frameworks for implementation, undertaking more detailed needs assessments as required (e.g. focus groups).

## CONCLUSION

This survey exploring medical students’ knowledge and perceptions of AI in medicine found an overall optimism on AI’s role in healthcare and enthusiasm to obtain AI competencies beginning in undergraduate medical training. As AI-enabled technologies are increasingly integrated into healthcare, medical education must adapt to continue producing competent physicians who can deliver high-quality patient care.

## Supporting information

Full Survey

## Data Availability

Full survey data is available upon request.

