## Supplemental Tables for "Knowledge of and Attitudes on Artificial Intelligence in Healthcare: A Provincial Survey Study of Medical Students"

**APPENDIX**

**Appendix Table 1: The perceived ability of AI to eventually perform a specific task at the level of a human physician in domains of individual health, health systems, and population health.**

| **Capability** | **Extremely Likely** | **Likely** | **Uncertain** | **Unlikely** | **Extremely Unlikely** |
| --- | --- | --- | --- | --- | --- |
| *Provide patients with preventative health recommendations (e.g. exercise, diet, wellness).* | 19.4% (58/299)) | 46.2% (138/299) | 14.7% (44/299) | 16.7% (50/299) | 3.0% (9/299) |
| *Analyze patient information to reach diagnoses.* | 17.5% (52/297) | 53.5% (159/297) | 19.5% (58/297) | 8.8% (26/297) | 0.7% (2/297) |
| *Analyze patient information to establish prognoses.* | 19.9% (59/296) | 56.4% (167/296) | 16.9% (50/296) | 6.4% (19/296) | 0.3% (1/296) |
| *Read and interpret diagnostic imaging.* | 43.2% (126/292) | 40.1% (117/292) | 9.6% (28/292) | 6.2% (18/292) | 1.0% (3/292) |
| *Evaluate when to refer patients to other health professionals.* | 11.0% (32/290) | 31.0% (90/290) | 32.8% (95/290) | 23.8% (69/290) | 1.4% (4/290) |
| *Formulate personalized treatment plans for patients.* | 6.5% (19/292) | 22.6% (66/292) | 33.2% (97/292) | 31.2% (91/292) | 6.5% (19/292) |
| *Formulate personalized medication prescriptions for patients.* | 12.0% (35/292) | 41.4% (121/292) | 29.5% (86/292) | 14.0% (41/292) | 3.1% (9/292) |
| *Provide empathetic care to patients.* | 3.4% (10/292) | 3.1% (9/292) | 15.1% (44/292) | 28.8% (84/292) | 49.7% (145/292) |
| *Monitor patient compliance to prescribed medications, exercise and dietary recommendations.* | 13.4% (39/292) | 42.8% (125/292) | 22.3% (65/292) | 15.8% (46/292) | 5.8% (17/292) |
| *Provide psychiatric/personal counselling.* | 1.7% (5/290) | 5.2% (15/290) | 16.2% (47/290) | 32.4% (94/290) | 44.5% (129/290) |
| *Perform surgery (e.g. robotic surgery).* | 15.2% (44/289) | 38.8% (112/289) | 24.2% (70/289) | 17.7% (51/289) | 4.2% (12/289) |
| *Provide documentation (e.g., update medical records) about patients.* | 38.8% (112/289) | 49.1% (142/289) | 8.3% (24/289) | 3.5% (10/289) | 0.4% (1/289) |
| *Assist hospitals in capacity planning and human resource management.* | 27.1% (78/288) | 45.5% (131/288) | 21.9% (63/288) | 4.2% (12/288) | 1.4% (4/288) |
| *Provide recommendations for quality improvement in practices/hospitals.* | 19.4% (56/289) | 45.7% (132/289) | 24.6% (71/289) | 9.7% (28/289) | 0.7% (2/289) |
| *Conduct population health surveillance and outbreak prevention.* | 29.4% (85/289) | 50.2% (145/289) | 15.9% (46/289) | 4.5% (13/289) | 0.0% (0/289) |
| *Select the best population health interventions.* | 12.1% (35/289) | 28.0% (81/289) | 40.8% (118/289) | 15.9% (46/289) | 3.1% (9/289) |

Shaded box = plurality opinion. Any response category within 5% of plurality opinion is also shaded.

**Appendix Table 2: Perceived timeline for AI to perform a specific task at the level of a human physician in domains of individual health, health systems, and population health.**

| **Capability*** | **0-4 years** | **5-10 years** | **11-25 years** | **26-50 years** | **> 50 years** |
| --- | --- | --- | --- | --- | --- |
| *Provide patients with preventative health recommendations (e.g. exercise, diet, wellness).* | 21.1% (41/194) | 31.4% (61/194) | 37.1% (72/194) | 6.7% (13/194) | 3.6% (7/194) |
| *Analyze patient information to reach diagnoses.* | 6.6% (14/213) | 29.6% (63/213) | 43.7% (93/213) | 16.4% (35/213) | 3.8% (8/213) |
| *Analyze patient information to establish prognoses.* | 13.3% (30/226) | 30.1% (68/226) | 44.3% (100/226) | 8.0% (18/226) | 4.4% (10/226) |
| *Read and interpret diagnostic imaging.* | 22.9% (56/245) | 44.1% (108/245) | 25.3% (62/245) | 5.3% (13/245) | 2.5% (6/245) |
| *Evaluate when to refer patients to other health professionals.* | 19.2% (24/125) | 35.2% (44/125) | 36.8% (46/125) | 5.6% (7/125) | 3.2% (4/125) |
| *Formulate personalized treatment plans for patients.* | 7.1% (6/85) | 24.7% (21/85) | 45.9% (39/85) | 15.3% (13/85) | 7.1% (6/85) |
| *Formulate personalized medication prescriptions for patients.* | 12.8% (20/156) | 35.9% (56/156) | 35.9% (56/156) | 13.5% (21/156) | 1.9% (3/156) |
| *Provide empathetic care to patients.* | 0.0% (0/20) | 25.0% (5/20) | 25.0% (5/20) | 20.0% (4/20) | 30.0% (6/20) |
| *Monitor patient compliance to prescribed medications, exercise and dietary recommendations.* | 22.7% (37/163) | 36.8% (60/163) | 31.3% (51/163) | 4.9% (8/163) | 4.3% (7/163) |
| *Provide psychiatric/personal counselling.* | 4.8% (1/20) | 33.3% (7/20) | 23.8% (5/20) | 19.1% (4/20) | 19.1% (4/20) |
| *Perform surgery (e.g. robotic surgery).* | 9.6% (15/156) | 24.4% (38/156) | 33.3% (52/156) | 21.8% (34/156) | 10.9% (17/156) |
| *Provide documentation (e.g. update medical records) about patients.* | 28.0% (71/254) | 41.7% (106/254) | 23.2% (59/254) | 6.30% (16/254) | 0.80% (2/254) |
| *Assist hospitals in capacity planning and human resource management.* | 25.7% (54/210) | 44.8% (94/210) | 23.8% (50/210) | 5.2% (11/210) | 0.5% (1/210) |
| *Provide recommendations for quality improvement in practices/hospitals.* | 24.6% (46/187) | 36.4% (68/187) | 29.4% (55/187) | 8.6% (16/187) | 1.1% (2/187) |
| *Conduct population health surveillance and outbreak prevention.* | 26.0% (60/231) | 38.5% (89/231) | 23.4% (54/231) | 9.5% (22/231) | 2.6% (6/231) |
| *Select the best population health interventions.* | 15.5% (18/116) | 43.1% (50/116) | 28.5% (33/116) | 12.1% (14/116) | 0.9% (1/116) |

*Only those who selected ‘extremely likely’ or ‘likely’ for AI to be able to eventually perform a specific capability were asked this question.

Shaded box = plurality opinion. Any response category within 5% of plurality opinion is also shaded.

**Appendix Table 3: Perceived impact of AI on medical profession, challenges raised by AI, and Integration of AI into the medical profession**

| **Prompt** | **Strongly agree** | **Agree** | **Neither** | **Disagree** | **Strongly disagree** |
| --- | --- | --- | --- | --- | --- |
| *Artificial Intelligence will reduce the number of jobs available to physicians.* | 5.2% (15/288) | 34.0% (98/288) | 33.7% (97/288) | 25.0% (72/288) | 2.1% (6/288) |
| *Artificial Intelligence will reduce the number of jobs in certain medical specialties more than others.* | 31.6% (91/288) | 55.2% (159/288) | 7.6% (22/288) | 4.7% (14/288) | 0.7% (2/288) |
| *Artificial Intelligence will/already did impact my choice of specialty selection.* | 3.1% (9/288) | 21.5% (62/288) | 17.0% (49/288) | 35.1% (101/288) | 23.3% (67/288) |
| *AI in medicine will raise new ethical challenges.* | 55.6% (160/288) | 42.0% (121/288) | 2.1% (6/288) | 0.4% (1/288) | 0.0% (0/288) |
| *AI in medicine will raise new social challenges.* | 50.7% (146/288) | 44.8% (126/288) | 3.5% (10/288) | 0.4% (1/288) | 0.7% (2/288) |
| *AI in medicine will raise new challenges around health equity.* | 41.3% (119/288) | 36.8% (106/288) | 16.0% (46/288) | 4.9% (14/288) | 1.0% (3/288) |
| *The Canadian healthcare system is currently well prepared to deal with challenges having to do with AI.* | 0.4% (1/288) | 3.8% (11/288) | 33.0% (95/288) | 47.6% (137/288) | 15.3% (44/288) |
| *My medical education is adequately preparing me for working alongside AI tools.* | 0.0% (0/288) | 2.4% (7/288) | 18.4% (53/288) | 53.1% (153/288) | 26.0% (75/288) |
| *Medical training should include training on AI competencies.* | 18.1% (52/287) | 54.0% (155/287) | 18.8% (54/287) | 7.3% (21/287) | 1.7% (5/287) |
| *Every medical trainee should be required to receive training in AI competencies.* | 11.9% (34/287) | 39.7% (114/287) | 27.2% (78/287) | 17.1% (49/287) | 4.2% (12/287) |
|  | **Medical student** | **Resident** | **Practicing Physician** | **None necessary** |  |
| *Training in AI competencies should begin as a:* | 68.2% (195/286) | 19.9% (57/286) | 1.1% (3/286) | 10.8% (31/286) |  |

Shaded box = plurality opinion. Any response category within 5% of plurality opinion is also shaded.

**Appendix Figure 1: Full survey questionnaire**

Seperately attached.
