## Supplementary material for "Knowledge of and Attitudes on Artificial Intelligence in Healthcare: A Provincial Survey Study of Medical Students": Full Survey

**Consent Page**

Agreement: Continuing to the next page (where you will begin the survey) indicates that you have read the information below, have had a chance to ask any questions you have about the study, and consent to participating in this study. 
-----------------------------------------------------------------------------------------

*Title of the study: Assessing Medical Trainees’ Knowledge and Perceptions of Artificial Intelligence in Medicine*

Invitation to Participate: You are invited to participate in the abovementioned research study conducted by Nishila Mehta, who is being supervised and advised by Dr. Sunit Das, Dr. Shiphra Ginsburg and Dr. Marcus Law. This project is supported by the Center of Ethics, University of Toronto. 

Participation: If you wish to participate in this study, please complete the survey. The survey should take you approximately 5-10 minutes to complete. You do not have to answer any questions that you do not want to answer, and may close the browser at anytime to be withdrawn from the study. 

Purpose of the Study: From this research we wish to learn assess medical students’ understanding of AI, and their perceptions around how AI will impact the healthcare field and their own careers. The results will inform our understanding of what existing gaps and opportunities there are for medical education to prepare future physicians for AI integrated healthcare environments.

Benefits: Respondents will have the option to enter into a raffle for a $25 Amazon giftcard. There is no other direct benefit from participating in this research study. 

Risks: There are no foreseeable risks.

Compensation: No compensation is provided.

Confidentiality and Anonymity: Your survey answers will be collected through SurveyMonkey, after which the data will be stored in a password protected electronic format. SurveyMonkey does not collect identifying information, therefore your responses will remain anonymous. 

The information that you will share will remain strictly confidential and will be used solely for the purposes of this research. The only people who will have access to the research data are the research student and principle investigator. Your answers to open-ended questions may be used verbatim in presentations and publications but you will not be identified. Results will be published in pooled (aggregate) format. 

The research study you are participating in may be reviewed for quality assurance to make sure that the required laws and guidelines are followed. If chosen, (a) representative(s) of the Human Research Ethics Program (HREP) may access study-related data and/or consent materials as part of the review. All information accessed by the HREP will be upheld to the same level of confidentiality that has been stated by the research team.

Conservation of data: The completed surveys will be stored in a password protected electronic format for a period of 5 years at which time they will be destroyed. 

Voluntary Participation: You are under no obligation to participate and if you choose to participate, you may withdraw at any time by closing your browser, or decline to answer any question(s) without any negative consequences.

Information about the Study Results: Study results will be available to study participants in the form of an academic publication. 

If you have any questions or require more information about the study itself, you may contact the researcher or his/her supervisor.

If you have any questions with regards to the ethical conduct of this study, you may contact the Research Oversight and Compliance Office - Human Research Ethics Program at or 416-946-3273.

**Section 1: Demographic and Background Information**

1. What medical school do you currently attend?
   a. University of Toronto
   b. Northern Ontario School of Medicine
   c. Western University
   d. Queen’s University
2. What year of medical school are you in?
   a. Year 1 (pre-clerkship)
   b. Year 2 (pre-clerkship)
   c. Year 3 (clerkship)
   d. Year 4 (clerkship)
   e. Other (please specify): _______________
3. What is your gender?
   a. Female
   b. Male
   c. Non-binary/ other gender
   d. Prefer not to say
   e. Prefer to self-describe: _________________
4. What did you do your undergraduate degree in? Please include the program name & degree obtained: __________________
5. If applicable, what did you do your Masters degree in? Please include the program name & degree obtained: _________________
6. If applicable, what did you do your PhD degree in? Please include the program name & degree obtained: _________________
7. Do you have an academic background in computer science?
   a. Yes
   b. No
8. Have you undertaken any additional training in computer science where artificial intelligence, machine learning or deep learning was taught/discussed (e.g. certificate program, online course, etc).
   a. Yes
      i. Please describe the training, including duration of the course(s): _______
   b. No
9. Have you attended or viewed any talks or lectures on artificial intelligence?
   a. Yes
      i. Approximately how many? ________
   b. No
10. Do you have any training in programming/coding?
    a. Yes
       i. Where did you acquire this training? ________
    b. No

**Section 2: Knowledge of Artificial Intelligence**

*To what extent do you agree with the following statements?*

1. I understand what the term “artificial intelligence” means.
   a. Strongly disagree
   b. Disagree
   c. Neither agree nor disagree
   d. Agree
   e. Strongly agree
2. I understand what the term “machine learning” means.
   a. Strongly disagree
   b. Disagree
   c. Neither agree nor disagree
   d. Agree
   e. Strongly agree
3. I understand what the term “neural network” means.
   a. Strongly disagree
   b. Disagree
   c. Neither agree nor disagree
   d. Agree
   e. Strongly agree
4. I understand what the term “deep learning” means.
   a. Strongly disagree
   b. Disagree
   c. Neither agree nor disagree
   d. Agree
   e. Strongly agree
5. I know what an algorithm is in the context of computer science.
   a. Strongly disagree
   b. Disagree
   c. Neither agree nor disagree
   d. Agree
   e. Strongly agree

**Section 3: Perceptions of Artificial Intelligence**

**Please read this definition:** Artificial Intelligence is the theory and development of computer systems able to perform tasks that normally require human intelligence, such as visual perception, speech recognition, and decision-making. 

In your opinion, **what is the likelihood that artificial intelligence enabled technologies will ever be able to replace human beings in performing these tasks** as well as or better than the average healthcare professional?

*Note: each time you select "extremely likely" or "likely", it will prompt a question regarding the timeline in which you see these innovations happening.

*Individual Patient Care*

1. Provide patients with preventative health recommendations (e.g. exercise, diet, wellness).
   a) Extremely unlikely
   b) Unlikely
   c) Likely
   d) Extremely likely
   e) Uncertain
      i. If answered Likely or Extremely likely: When, in your estimation, will technology have the capacity to replace the average healthcare professional in performing this task?
         - 0-4 years
         - 5 to 10 years from now
         - 11 to 25 years from now
         - 26-50 years
         - more than 50 years from now
2. Analyze patient information to reach diagnoses.
   a) Extremely unlikely
   b) Unlikely
   c) Likely
   d) Extremely likely
   e) Uncertain
      i. If answered Likely or Extremely likely: When, in your estimation, will technology have the capacity to replace the average healthcare professional in performing this task?
         - 0-4 years
         - 5 to 10 years from now
         - 11 to 25 years from now
         - 26-50 years
         - more than 50 years from now
3. Analyze patient information to establish prognoses.
   a) Extremely unlikely
   b) Unlikely
   c) Likely
   d) Extremely likely
   e) Uncertain
      i. If answered Likely or Extremely likely: When, in your estimation, will technology have the capacity to replace the average healthcare professional in performing this task?
         - 0-4 years
         - 5 to 10 years from now
         - 11 to 25 years from now
         - 26-50 years
         - more than 50 years from now
4. Read and interpret diagnostic imaging.
   a) Extremely unlikely
   b) Unlikely
   c) Likely
   d) Extremely likely
   e) Uncertain
      i. If answered Likely or Extremely likely: When, in your estimation, will technology have the capacity to replace the average healthcare professional in performing this task?
         - 0-4 years
         - 5 to 10 years from now
         - 11 to 25 years from now
         - 26-50 years
         - more than 50 years from now
5. Evaluate when to refer patients to other health professionals.
   a) Extremely unlikely
   b) Unlikely
   c) Likely
   d) Extremely likely
   e) Uncertain
      i. If answered Likely or Extremely likely: When, in your estimation, will technology have the capacity to replace the average healthcare professional in performing this task?
         - 0-4 years
         - 5 to 10 years from now
         - 11 to 25 years from now
         - 26-50 years
         - more than 50 years from now
6. Formulate personalized treatment plans for patients.
   a) Extremely unlikely
   b) Unlikely
   c) Likely
   d) Extremely likely
   e) Uncertain
      i. If answered Likely or Extremely likely: When, in your estimation, will technology have the capacity to replace the average healthcare professional in performing this task?
         - 0-4 years
         - 5 to 10 years from now
         - 11 to 25 years from now
         - 26-50 years
         - more than 50 years from now
7. Formulate personalized medication prescriptions for patients.
   a) Extremely unlikely
   b) Unlikely
   c) Likely
   d) Extremely likely
   e) Uncertain
      i. If answered Likely or Extremely likely: When, in your estimation, will technology have the capacity to replace the average healthcare professional in performing this task?
         - 0-4 years
         - 5 to 10 years from now
         - 11 to 25 years from now
         - 26-50 years
         - more than 50 years from now
8. Provide empathetic care to patients.
   a) Extremely unlikely
   b) Unlikely
   c) Likely
   d) Extremely likely
   e) Uncertain
      i. If answered Likely or Extremely likely: When, in your estimation, will technology have the capacity to replace the average healthcare professional in performing this task?
         - 0-4 years
         - 5 to 10 years from now
         - 11 to 25 years from now
         - 26-50 years
         - more than 50 years from now
9. Monitor patient compliance to prescribed medications, exercise and dietary recommendations.
   a) Extremely unlikely
   b) Unlikely
   c) Likely
   d) Extremely likely
   e) Uncertain
      i. If answered Likely or Extremely likely: When, in your estimation, will technology have the capacity to replace the average healthcare professional in performing this task?
         - 0-4 years
         - 5 to 10 years from now
         - 11 to 25 years from now
         - 26-50 years
         - more than 50 years from now
10. Provide psychiatric/personal counselling.
    a) Extremely unlikely
    b) Unlikely
    c) Likely
    d) Extremely likely
    e) Uncertain
       i. If answered Likely or Extremely likely: When, in your estimation, will technology have the capacity to replace the average healthcare professional in performing this task?
          - 0-4 years
          - 5 to 10 years from now
          - 11 to 25 years from now
          - 26-50 years
          - more than 50 years from now
11. Perform surgery (e.g. robotic surgery).
    a) Extremely unlikely
    b) Unlikely
    c) Likely
    d) Extremely likely
    e) Uncertain
       i. If answered Likely or Extremely likely: When, in your estimation, will technology have the capacity to replace the average healthcare professional in performing this task?
          - 0-4 years
          - 5 to 10 years from now
          - 11 to 25 years from now
          - 26-50 years
          - more than 50 years from now

*Health Systems*

12. Provide documentation (e.g., update medical records) about patients.
    a) Extremely unlikely
    b) Unlikely
    c) Likely
    d) Extremely likely
    e) Uncertain
       i. If answered Likely or Extremely likely: When, in your estimation, will technology have the capacity to replace the average healthcare professional in performing this task?
          - 0-4 years
          - 5 to 10 years from now
          - 11 to 25 years from now
          - 26-50 years
          - more than 50 years from now
13. Assist hospitals in capacity planning and human resource management.
    a) Extremely unlikely
    b) Unlikely
    c) Likely
    d) Extremely likely
    e) Uncertain
       i. If answered Likely or Extremely likely: When, in your estimation, will technology have the capacity to replace the average healthcare professional in performing this task?
          - 0-4 years
          - 5 to 10 years from now
          - 11 to 25 years from now
          - 26-50 years
          - more than 50 years from now
14. Provide recommendations for quality improvement in practices/hospitals.
    a) Extremely unlikely
    b) Unlikely
    c) Likely
    d) Extremely likely
    e) Uncertain
       i. If answered Likely or Extremely likely: When, in your estimation, will technology have the capacity to replace the average healthcare professional in performing this task?
          - 0-4 years
          - 5 to 10 years from now
          - 11 to 25 years from now
          - 26-50 years
          - more than 50 years from now

*Population Health*

15. Conduct population health surveillance and outbreak prevention.
    a) Extremely unlikely
    b) Unlikely
    c) Likely
    d) Extremely likely
    e) Uncertain
       i. If answered Likely or Extremely likely: When, in your estimation, will technology have the capacity to replace the average healthcare professional in performing this task?
          - 0-4 years
          - 5 to 10 years from now
          - 11 to 25 years from now
          - 26-50 years
          - more than 50 years from now
16. Select the best population health interventions.
    a) Extremely unlikely
    b) Unlikely
    c) Likely
    d) Extremely likely
    e) Uncertain
       i. If answered Likely or Extremely likely: When, in your estimation, will technology have the capacity to replace the average healthcare professional in performing this task?
          - 0-4 years
          - 5 to 10 years from now
          - 11 to 25 years from now
          - 26-50 years
          - more than 50 years from now

*Impact on the Medical Profession*

17. Artificial Intelligence will reduce the number of jobs available to me.
    a) Strongly agree
    b) Agree
    c) Neither agree nor disagree
    d) Disagree
    e) Strongly disagree
18. Artificial Intelligence will reduce the number of jobs in certain medical specialties more than others.
    a) Strongly agree
    b) Agree
    c) Neither agree nor disagree
    d) Disagree
    e) Strongly disagree
19. Artificial Intelligence will/already did impact my choice of specialty selection.
    a) Strongly agree
    b) Agree
    c) Neither agree nor disagree
    d) Disagree
    e) Strongly disagree

*AI Ethics*

20. AI in medicine will raise new ethical challenges.
    a) Strongly agree
    b) Agree
    c) Neither agree nor disagree
    d) Disagree
    e) Strongly disagree
21. AI in medicine will raise new social challenges.
    a) Strongly agree
    b) Agree
    c) Neither agree nor disagree
    d) Disagree
    e) Strongly disagree
22. AI in medicine will raise new challenges around health equity.
    a) Strongly agree
    b) Agree
    c) Neither agree nor disagree
    d) Disagree
    e) Strongly disagree
23. The Canadian healthcare system is currently well prepared to deal with challenges having to do with AI.
    a) Strongly agree
    b) Agree
    c) Neither agree nor disagree
    d) Disagree
    e) Strongly disagree

*AI and medical education*

24. My medical education is adequately preparing me for working alongside AI tools
    a) Strongly agree
    b) Agree
    c) Neither agree nor disagree
    d) Disagree
    e) Strongly disagree
25. Medical training should include training on AI competencies (e.g. what is AI, how will it impact us, what are the challenges it raises).
    a) Strongly agree
    b) Agree
    c) Neither agree nor disagree
    d) Disagree
    e) Strongly disagree
26. Every medical trainee should be required to receive training in AI competencies.
    a) Strongly agree
    b) Agree
    c) Neither agree nor disagree
    d) Disagree
    e) Strongly disagree
27. Training in AI competencies should begin as a:
    a) Medical Student
    b) Resident
    c) Practicing Physician
    d) No training in AI competencies is necessary
28. Do you have any comments or concerns on the topic of AI in medicine?
